# ^18^F-FDG PET/CT characteristics of IASLC grade 3 invasive adenocarcinoma and the value of ^18^F-FDG PET/CT for preoperative prediction

**DOI:** 10.1101/2023.10.04.23296555

**Authors:** Hanyun Yang, Xinran Liu, Lijuan Wang, Wenlan Zhou, Ying Tian, Ye Dong, Kemin Zhou, Li Chen, Meng Wang, Hubing Wu

## Abstract

**Purpose:** This study is performed to investigate the imaging characteristics of the International Association for the study of lung cancer (IASLC) grade 3 invasive adenocarcinoma (IAC) on PET/CT and the value of PET/CT for preoperative predicting this tumor.

**Materials and Methods:** We retrospectively enrolled patients with IAC from August 2015 to September 2022. The clinical characteristics, serum tumor markers, and PET/CT features were analyzed. T test, Mann-Whitney U test, χ^2^ test, Logistic regression analysis, and receiver operating characteristic (ROC) analysis were used to predict grade 3 tumor and evaluate the prediction effectiveness.

**Results:** Grade 3 tumors had a significantly higher maximum standardized uptake value (SUV_max_) (*P* < 0.001), while Grade 1 - 2 tumors were prone to present with air bronchogram sign or vacuole sign (*P* < 0.001). Multivariate logistic regression analysis revealed that only SUV_max_ (OR = 1.137; 95% CI: 1.037, 1.247; *P* < 0.05) and air bronchogram sign or vacuole sign (OR = 0.225; 95% CI: 0.088, 0.572; *P* < 0.05) were independent predictors for Grade 3 tumors. The established prediction formula for Grade 3 tumors was P = one / [one + EXP (1.112 - 0.187 × SUV_max_ + 1.395 × air bronchogram sign or vacuole sign)], which generated a high AUC (0.825) and negative predictive value (0.924), respectively.

**Conclusion:** Our study demonstrates that grade 3 IAC has a unique PET/CT imaging feature. The prediction model established with SUV_max_ and air bronchogram sign or vacuole sign can effectively predict grade 3 tumors before the operation.

## Introduction

Lung cancer is the most common malignant tumor globally, especially in China, and has a relatively poor prognosis.^1–3^ In the different pathological types of lung cancer, lung adenocarcinoma is the most heterogeneous tumor, which can be further divided into non-invasive adenocarcinoma and invasive adenocarcinoma (IAC).^4,5^ In 2020, the International Association for the study of lung cancer (IASLC) proposed a new invasive lung adenocarcinoma grading system (IASLC grading system), which emphasized the role of high-grade pattern on grading IAC, differed from lung adenocarcinoma grading system 2015.^6,7^ According to IASLC grading 2020 system, the IAC was classified into three grades: grade 1, lepidic predominant tumor; grade 2, acinar or papillary predominant tumor, both with no or less than 20% of high-grade patterns; and grade 3, any tumor with 20% or more of high-grade patterns (solid, micropapillary, or complex gland).^6,7^ Since new IASLC grading system was proposed, more and more studies have confirmed that the new grading system can represent the tumor aggressiveness and predict the prognosis better than 2015 version.^8–10^ Among the different subtypes of IAC, IASLC grade 3 tumor is reported to have a higher incidence of mediastinal lymph node metastasis^11^, a much lower five - year survival rate than grade 1 or 2 and can serve as an independent predictor of poor prognosis of IAC.^12,13^ For most grade 1 or 2 tumors, the lesions can be treated only by surgery and the patients always have good prognosis,^14^ however, for IASLC grade 3 tumor, preoperative neoadjuvant chemoradiotherapy and postoperative adjuvant chemoradiotherapy may be benefit for the patients to achieve a good prognosis.^3,15,16^ More aggressive surgery, such as lobectomy and systematic mediastinal lymph node resection, is also needed for these tumors.^17,18^ Due to the high aggressiveness of this tumor, it is of great clinical value to predict it noninvasively before the surgery, which is useful to guide surgeon to make a more suitable treatment plan.

^18^F-fluorodeoxyglucose (^18^F-FDG) positron emission tomography/computed tomography (PET/CT) is an important modality for tumor imaging, which can not only provide the detailed morphologic changes to represent the pathological process of the lesions through CT component, but also can provide the tumor’s metabolic activity through PET component. The tumor’s metabolic activity was always reported to be closely related to tumor aggressiveness and the prognosis of the patients.^19–21^ Currently, PET/CT has gained a great application on the diagnosis, pretherapeutic staging and efficacy evaluation of lung cancer, ^22–24^ however, few studies were reported concerning its use in evaluating the tumor grade based on the new IASLC grading system 2020. In the present study, we retrospectively analyzed the ^18^F-FDG PET/CT findings in 227 patients to uncover the imaging characteristics of IASLC grade 3 tumor and investigate the value of ^18^F-FDG PET/CT for noninvasively predicting IASLC grade 3 tumors before surgery.

## Materials and methods

### Study subjects

A totality of 227 IAC, patients who underwent ^18^F-FDG PET/CT scan in Nanfang Hospital from August 2015 to September 2022, were enrolled in this retrospectively study. The inclusion criteria were as follows: (1) Patients who did not receive any anti-tumor treatment before PET/CT scan; (2) Primary IAC was diagnosed by pathology and various tissue pathological components (such as adherent, acinar, nipple, or high - grade components) and their proportion could be identified; (3) Patients with early tumor stage and received the surgical resection for the tumor. The exclusion criteria were: (1) Patients had previous history of other tumors; (2) Although IAC was diagnosed, the tissue pathological component and their proportion were not clearly identified; (3) Diabetic patients with fasting blood glucose ≥ 11.1 mmol/L; (4) Patients had not received surgical resection for the tumor; (5) Patients with or mixed with other pathological types of lung cancer, such as squamous cell carcinoma or small cell carcinoma.

### ^18^F-FDG PET/CT acquisition

^18^F-FDG was automatically synthesized using a PET trace cyclotron (GE Healthcare) and the ^18^F-FDG synthesizer module Tracerlab FXF-N (Beijing PET Biotechnology Co. Ltd). PET/CT images were acquired using Biograph mCTx (Siemens, Germany) for 151 patients and using uEXPLORER PET/CT (United Imaging Corporation, China) for other 76 patients.^25^ The detailed ^18^F-FDG PET/CT acquisition see the supplementary materials (1).

### Thin-section acquisition

After completion of whole-body PET/CT scan, a limited thin-section CT was performed for the lung nodules. The detailed thin-section acquisition was provided in the supplementary materials (2).

### PET/CT and thin-section CT image interpretation

All PET/CT images were independently evaluated by two experienced PET/CT physicians. On PET/CT, the lung nodule was identified on CT images and PET metabolic parameters, the maximum standardized uptake value (SUV_max_), metabolic tumor volume (MTV), and total lesion glycolysis (TLG), were measured on PET. To obtained these parameters, a region of interest (ROI) covering the lesion on the axial PET/CT image was drawn, and automatically changed to a 3D volume of interest under the 60% isovolumetric line. A 40% SUV_max_ threshold method was used to define the ROI, and SUV_max_, MTV, and TLG were then automatically measured.

When the thin-section CT images were analyzed, the width of the lung window was set at 1200 Hu, and the window level was −600 Hu; while the width of the mediastinal window was 300 Hu, and the window level was 40 Hu. On CT images, the CT features, such as lobulated sign, burr sign, vascular sign, air containing bronchus sign or vacuole sign, pleural traction sign, and the tumor attenuation [solid nodule, mixed nodule or pure ground glass nodule (pGGN)], were identified by two experienced radiologists. The size of the lesion [expressed as the maximum diameter of the lesion (cm)], and consolidation-tumor-ratio (CTR) were measured according to the published literatures.^26,27^

### Tumor markers

Tumor marker, such as serum cytokeratin fragment 21 - 1 (CYFRA21 - 1), carcinoembryonic antigen (CEA), squamous cell carcinoma antigen (SCC), neuron-specific enolase (NSE), were measured by standard essay method by the clinical laboratory of our hospital, and the data could be obtained through the hospital picture archiving and communications system (PACS).

### Pathological examination

All the samples for pathological examination were obtained by surgery. The pathological diagnosis and the detailed information, such as the tissue pathological components and their proportion of the lesion were also obtained through the hospital PACS system. The grades for the tumors were re-classified according to the new grading system in 2020 by two experienced pathologists.^6^

### Statistical analysis

SPSS 26.0 statistical software was used to conduct univariate analysis on the case data. Continuous variables in line with normal distribution were expressed in x̄ ± s, and were tested the significance of the difference using independent samples *t* test. Continuous variables that did not conform to normal distribution were expressed in M (Q25, Q75), and Mann-Whitney U was used to test the significance of the difference. Count data were expressed by rate, using χ^2^ to test and to compare the difference of constituent ratio between groups. Logistic multiple regression analysis was used for multivariate analysis. Factors with *P* < 0.05 in univariate analysis and factors that clinical considered may be associated with IACs were included in logistic regression. ROC curve was used to evaluate the diagnostic efficiency of multiple regression model. Double sided inspection, ɑ = 0.05.

## Results

### Comparison of clinical characteristics between IASLC grade 3 and grade 1 or 2 IACs

227 patients with IAC (119 males, 108 females; mean age, 61.07 ± 9.12) from August 2015 to September 2022 were enrolled. According to the new IASLC grading system, 167 patients were diagnosed as grade 1 or 2 tumor (73.6%), and 60 were diagnosed as grade 3 tumor (26.4%). Biograph mCTx acquired 43 grade 1, 67 grade 2 and 41 grade 3 and uEXPLORER PET/CT acquired 19 grade 1, 38 grade 2 and 19 grade 3. Among 60 patients with grade 3 tumors, 10 (16.7%) were diagnosed to have mediastinal lymph node metastasis, which was significantly higher than that (11/167, 6.6%) of grade 1 or 2 tumors (χ^2^ = 5.342, *P* = 0.021). The stages of 167 patients with grade 1 or 2 tumor were 147 stage I, 8 stage II and 12 stage III, while the stages of 60 patients with grade 3 tumor were 42 stage I, 8 stage II and 10 stage III. The patient with male, smoking-history and older in age was more prone to suffer from IASLC grade 3 tumor (all *P* < 0.05, Table 1). Meanwhile, higher levels of serum CYFRA21 - 1, CEA and SCC were observed in the patients with IASLC grade 3 tumor than those in the patients with IASLC grade 1 or 2 tumors (all *P* < 0.05, Table 1).

**Table 1.** Univariate analysis of the relationship between the factors of clinical characteristics, tumor markers and ^18^F-FDG PET/CT features and IASLC grade 3 tumor.

| Factor | Grade 1 or 2 tumors<br>(n = 167) | Grade 3 tumors (n = 60) | $\chi^2/t/U$ | P value |
| --- | --- | --- | --- | --- |
| clinical characteristics |  |  |  |  |
| Sex<br>(male/female) | 80/87 | 39/21 | 5.173 | 0.023* |
| Age ($\bar{x} \pm s$) | 60.21 ± 8.95 | 63.47 ± 9.29 | -2.393 | 0.018* |
| Smoking history<br>(yes/no) | 56/111 | 35/25 | 11.304 | 0.001* |
| Tumor markers |  |  |  |  |
| CYFRA21 - 1<br>[M (Q<sub>25</sub>, Q<sub>75</sub>)] | 2.37 (1.73, 3.23) | 3.38 (2.26, 4.29) | 3256.0 | < 0.001* |
| CEA<br>[M (Q<sub>25</sub>, Q<sub>75</sub>)] | 1.56 (0.92, 2.58) | 3.54 (1.63, 6.50) | 2906.5 | < 0.001* |
| SCC<br>[M (Q<sub>25</sub>, Q<sub>75</sub>)] | 1.12 (0.75, 1.61) | 1.3(0.96, 1.95) | 4131.5 | 0.044* |
| NSE<br>[M (Q<sub>25</sub>, Q<sub>75</sub>)] | 12.3 (10.66, 14.35) | 12.26 (10.81, 14.01) | 4831.0 | 0.825 |
| CT features |  |  |  |  |
| Maximum diameter of lesion<br>[M (Q<sub>25</sub>, Q<sub>75</sub>)] | 1.8 (1.5, 2.4) | 1.9 (1.4, 2.5) | 4998.5 | 0.979 |
| CTR<br>[M (Q<sub>25</sub>, Q<sub>75</sub>)] | 0.75 (0.50, 1.00) | 1.00 (1.00, 1.00) | 2379.5 | < 0.001* |
| Solid tumor<br>(yes/no) | 50/117 | 49/11 | 48.026 | < 0.001* |
| Lobulated sign<br>(with/without) | 152/15 | 58/2 | 2.033 | 0.154 |
| Burr sign<br>(with/without) | 69/98 | 38/22 | 8.586 | 0.003* |
| Vascular sign<br>(with/without) | 128/39 | 49/11 | 0.648 | 0.421 |
| Air bronchogram sign or<br>vacuole sign<br>(with/without) | 149/18 | 31/29 | 37.919 | < 0.001* |
| Pleural traction sign<br>(with/without) | 117/50 | 47/13 | 1.507 | 0.220 |
| PET signs |  |  |  |  |
| SUV<sub>max</sub><br>[M (Q<sub>25</sub>, Q<sub>75</sub>)] | 2.50 (1.30, 5.10) | 8.61 (4.71, 12.90) | 2077.5 | < 0.001* |
| MTV<br>[M (Q<sub>25</sub>, Q<sub>75</sub>)] | 2.07 (1.16, 3.73) | 1.73 (0.82, 3.74) | 4867.0 | 0.743 |
| TLG<br>[M (Q<sub>25</sub>, Q<sub>75</sub>)] | 3.39 (1.62, 7.27) | 6.82 (3.95, 23.30) | 3146.0 | < 0.001* |

<sup>a</sup> CYFRA21 - 1, cytokeratin fragment 21 - 1; CEA, carcinoembryonic antigen; SCC, squamous cell carcinoma antigen; NSE, neuron-specific enolase; CTR, consolidation-tumor-ratio; SUV<sub>max</sub>, maximum standardized uptake value; MTV, metabolic tumor volume; TLG, total lesion glycolysis; CT, computed tomography; PET, positron emission tomography; M (Q<sub>25</sub>, Q<sub>75</sub>), median (25% quantile, 75% quantile).
\*Significant difference.

### ^18^F-FDG PET / CT imaging characteristics of IASLC grade 3 and grade 1 or 2 IACs

On CT images, grade 3 tumors had a similar size of lesion in the diameter to that of grade 1 or 2 tumors (*P* > 0.05, Table 1). There was also no significant difference in the incidence of lobulation sign, vascular sign and pleural traction sign between grade 3 and grade 1 or 2 tumors (all *P* > 0.05, Table 1). However, almost 80% (49/60) grade 3 tumors presented as solid lesions, and they had a higher CTR than that of grade 1 or 2 tumors (1.0 *vs*. 0.75, U = 2379.5, *P* < 0.001, Table 1). Meanwhile, burr sign was more commonly seen in grade 3 tumors compared to grade 1 or 2 tumors (63.3% *vs.* 41.3%, χ^2^ = 0.586, *P* = 0.003, Table1). On the contrary, the air bronchogram sign or vacuole sign (Fig. 1) was more commonly found in grade 1 or 2 tumors compared to grade 3 tumors (89.2% *vs.* 51.7%, χ^2^ = 37.919, *P* < 0.001, Table 1).

**Fig. 1.**
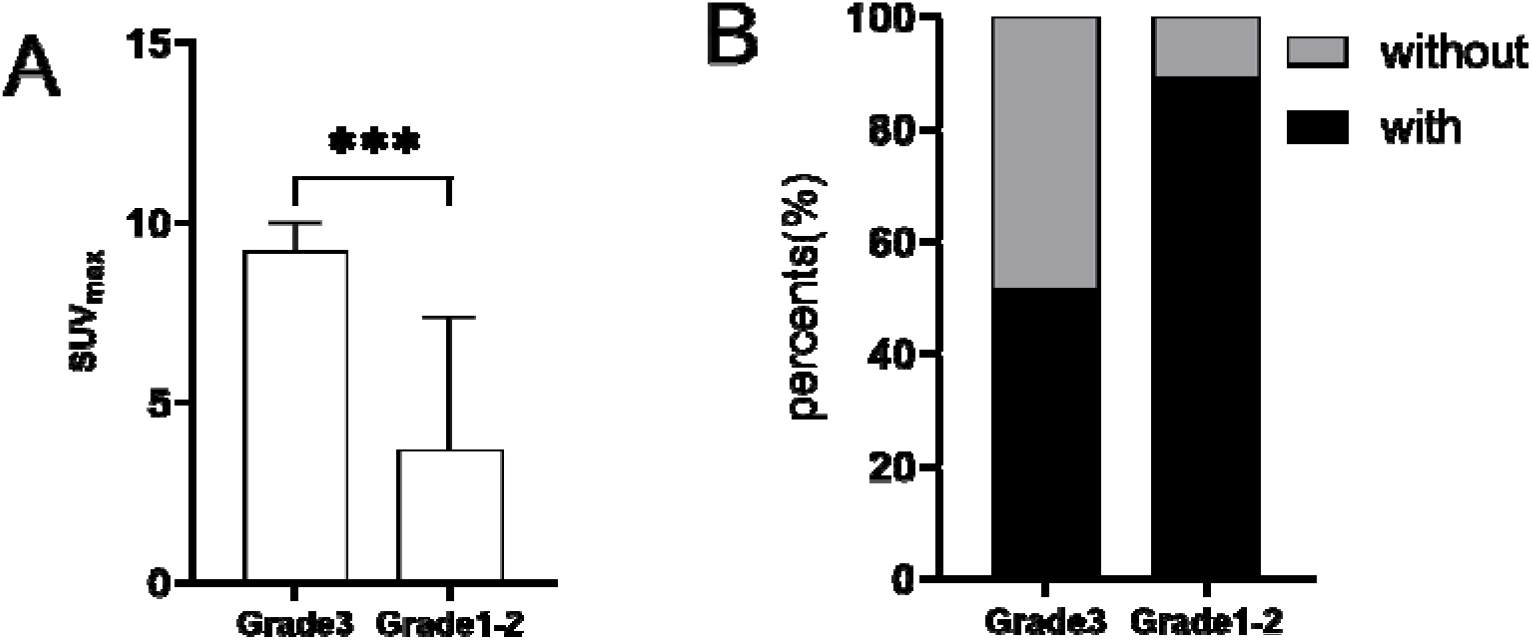
The SUV_max_ and the incident percent of air bronchogram sign or vacuole sign between IASLC grade 3 tumors and grade 1 or 2 tumors. A: SUV_max_; B: Incident percent of air bronchogram sign or vacuole sign. ^a^ SUV_max_, maximum standardized uptake value.

On PET images, ^18^F-FDG uptake higher than that of mediastinal blood pool was observed in 86.7% of the grade 3 tumors, however, only in 46.1% of the grade 1 or 2 tumors and the difference between them was significantly (86.7% *vs.* 46.1%, χ^2^ = 29.597, *P <* 0.001). Correspondingly, the median SUV_max_ of grade 3 tumors was observed to be significantly higher than that of grade 1 or 2 tumors [8.61 (4.71, 12.90) *vs.* 2.50 (1.30, 5.10), U = 2077.5, *P <* 0.001, Table 1]. Similarly, the median TLG of grade 3 tumors was also significantly higher than that of grade 1 or 2 tumors [6.82 (3.95, 23.30) *vs.* 3.39 (1.62, 7.27), U = 3146.0, *P <* 0.001, Table 1].

Typical ^18^F-FDG PET/CT images of grade 3 tumor and grade 1 or 2 tumors were shown in Fig. 2.

**Fig. 2.**
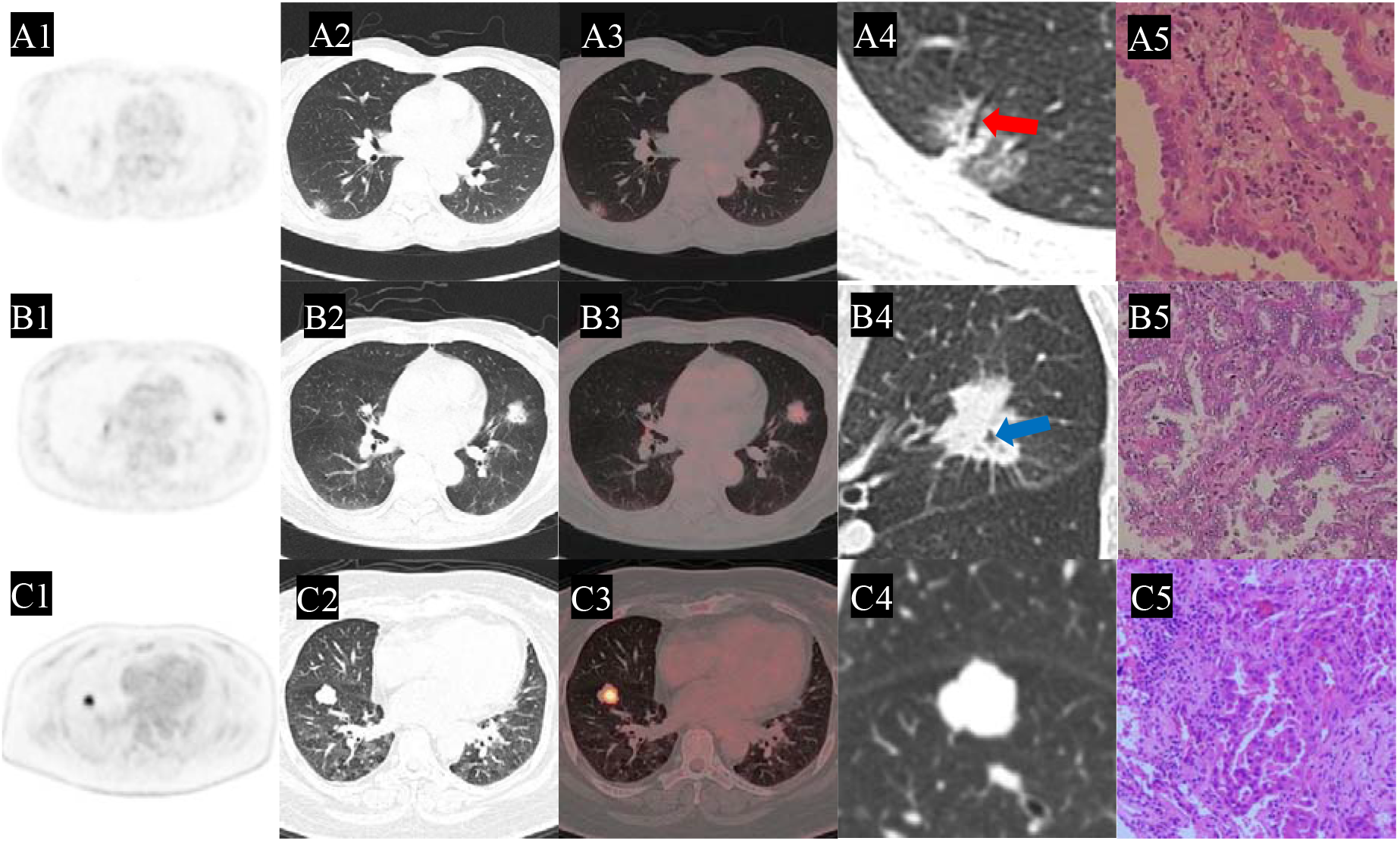
Typical ^18^F-FDG PET/CT imaging features of IASLC grade 3 tumor and 1or 2 tumor. A1 - A5: A patient suffered from IASLC grade 1 tumor. The pictures were PET, CT, PET/CT fusion, thin-section CT, and histopathological (HE staining, 400x) images, respectively. A mixed ground-glass nodule was detected in lower lobe of the right lung (A2), which had a mild uptake of ^18^F-FDG (SUV_max_ was 2.4, TLG was 2.1) (A3) and had an air bronchogram sign (A4). B1 - B5: A patient suffered from IASLC grade 2 tumor.The pictures were PET, CT, PET/CT fusion, thin-section CT, and histopathological (HE staining, 200x) images, respectively. A solid nodule was found in upper lobe of the left lung (B2), which had a medium uptake of ^18^F-FDG (SUV_max_ was 4.7, TLG was 9.7) (B3) and had an vacuole sign (B4). C1 - C5: A patient suffered from IASLC grade 3 tumor.The pictures were PET, CT, PET/CT fusion, thin-section CT, and histopathological (HE staining, 400x) images, respectively. A solid nodule was detected in lower lobe of the right lung (C2), which had an intense uptake of ^18^F-FDG (SUV_max_ was 9.6, TLG was 14.2) (C3) and no air bronchogram sign or vacuole sign was observed (C4). The red arrow points to air bronchogram sign. The blue arrow points to vacuole sign.

### Analysis of related factors to grade 3 invasive lung adenocarcinoma

A totality of 18 parameters including clinical characteristics, CT features and PET metabolic parameters were included in the univariate analysis to analyze the relationship of these parameters with tumor grades. It revealed that 12 parameters including sex, age, smoking history, CYFRA21 - 1, CEA, SCC, burr sign, air bronchogram sign or vacuole sign, CTR, solid tumor, SUV_max_ and TLG were closely related to tumor grades (all *P* < 0.05, Table 1), and other parameters (NSE, maximum diameter of lesion, lobulation sign, vascular sign, pleural traction sign, MTV) were not related to tumor grades (all *P* ≥ 0.05, Table 1). The above parameters related to tumor grade found in univariate analysis were further included into the multivariate logistic regression analysis. The results showed that only SUV_max_ (OR = 1.137, 95% confidence interval: 1.037 - 1.247, *P* < 0.05) and air bronchogram sign or vacuole sign (OR = 0.225, 95% confidence interval: 0.088 - 0.572, *P* < 0.05) were closely associated with tumor grade (all *P* < 0.05, Table 2).

**Table 2.** The independent factors for predicting IASLC grade 3 tumor determined using Multivariate analysis.

| Factor | *P* value | OR | 95% confidence interval |  |
| --- | --- | --- | --- | --- |
|  |  |  | Lower limit | Upper limit |
| Sex | 0.613 | 0.694 | 0.168 | 2.867 |
| Age | 0.345 | 1.024 | 0.975 | 1.075 |
| Smoking history | 0.082 | 3.512 | 0.852 | 14.474 |
| CYFRA21-1 | 0.086 | 1.243 | 0.969 | 1.594 |
| CEA | 0.077 | 1.144 | 0.985 | 1.327 |
| SCC | 0.661 | 0.924 | 0.648 | 1.317 |
| SUV<sub>max</sub> | 0.015* | 1.123 | 1.023 | 1.234 |
| TLG | 0.830 | 0.998 | 0.984 | 1.013 |
| Burr sign | 0.995 | 0.997 | 0.429 | 2.321 |
| Air bronchogram sign or vacuole sign | 0.006* | 0.259 | 0.099 | 0.678 |
| CTR | 0.168 | 5.586 | 0.485 | 64.339 |
| Solid tumor or not | 0.902 | 1.083 | 0.301 | 3.897 |
| Constant | 0.004 | 0.006 | / | / |

<sup>a</sup> CYFRA21 - 1, cytokeratin fragment 21 - 1; CEA, carcinoembryonic antigen; SCC, squamous cell carcinoma antigen; SUV<sub>max</sub>, maximum standardized uptake value; TLG, total lesion glycolysis; CTR, consolidation-tumor-ratio; OR, odds ratio.
\*Significant difference.

### Evaluation of ^18^F-FDG PET/CT for preoperative prediction of grade 3 IACs

The parameters (SUV_max_ and air bronchogram sign or vacuole sign) that were clearly related to tumor grades on multivariate analysis were selected into the logistic regression analysis, and the regression model formula for preoperative prediction of grade 3 IAC was established as: P = 1/(1 + e^−X^), where X = −1.112 + 0.187 × SUV_max_ - 1.395 × air bronchogram sign or vacuole sign. SUV_max_ should be given as the measured value, while air bronchogram sign or vacuole sign was entered with positive (default value = 1) and negative (default value = 0). The goodness-of-fit of the model was Nagelkerke R^2^ = 0.354, and the Hosmer - lemeshow test of the binary logistic regression showed χ^2^ = 11.142, and *P* = 0.194, which indicated a good fit (Table 3). The ROC curve of this predicting model was shown in Fig. 3, and the area under the curve (AUC) value was 0.825 (95% CI: 0.760 - 0.891), which showed a better prediction efficiency using SUV_max_ alone (AUC = 0.793, cut-off value = 3.504). The optimal cut-off value was *P* = 0.1849345, which prompts that the greater the *P* value is, the more likely the lesion is grade 3 tumor. The model generated the sensitivity, specificity, accuracy, positive predictive value and negative predictive value for preoperative prediction of grade 3 IAC were 0.833 (50/60), 0.725 (121/167), 0.753 (171/227), 0.521 (50/96), and 0.924 (121/131), respectively.

**Table 3.** Binary logistic regression results of a model for preparative predicting IASLC grade 3 tumor.

| Factor | B | P value | OR | 95% confidence interval |  |
| --- | --- | --- | --- | --- | --- |
|  |  |  |  | Lower limit | Upper limit |
| SUV<sub>max</sub> | 0.187 | < 0.001 | 1.206 | 1.118 | 1.301 |
| Air bronchogram sign or vacuole sign | -1.395 | 0.001 | 0.248 | 0.113 | 0.545 |
| Constant | -1.112 | 0.013 | 0.329 | / | / |

SUV<sub>max</sub>, maximum standardized uptake value; B, regression coefficient; OR, odds ratio.

**Fig. 3.**
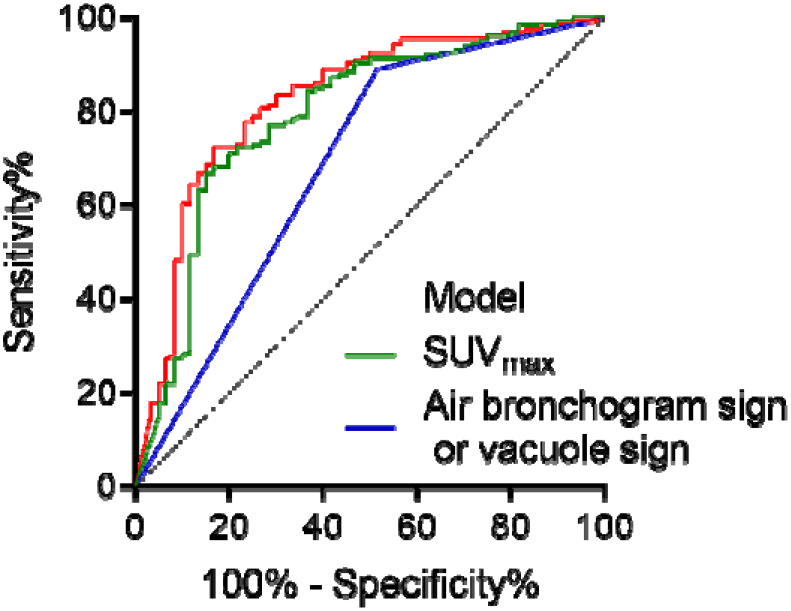
ROC curve of the model built on SUV_max_ and air bronchogram sign or vacuole sign for preoperative predicting IASLC grade grade 3 tumor.

## Discussion

Several studies had confirmed that IASLC grade 3 tumor actually had a higher aggressiveness. Long et al. reported that IASLC 3 grade tumors had a much higher mediastinal pN_2_ lymph node metastasis (18.3%) compared to 9.6% of grade 2 tumors and 1.4% of grade 1 tumors (*P* < 0.001).^11^ Our results showed a similar result that IASLC grade 3 tumors had mediastinal lymph node metastasis in 16.7% patients, which was significantly higher than 6.6% of grade 1 or 2 tumors (*P* = 0.021).

Accurate prediction of IASLC grade 3 tumors was of great significance to guide preoperative treatment and surgical resection. As a highly malignant tumor, many studies had shown that IASLC grade 3 tumors had relatively unique clinical and imaging characteristics. The studies of Ryo and Wongi showed that males, younger patients and smokers were more likely to suffer from IASLC grade 3 tumors.^28^ Different from Ryo’s study, our study found that more IASLC grade 3 tumors occurred in older patients. The biological characteristics of tumors might be more easily exhibited on imaging modalities. The study of Min et al. showed that CTR and tumor size were independent influencing factors for predicting high-grade IASLC. They found that CTR (cut-off values < 25% and ≥ 75%) and tumor size (cut-off value 17 mm) were helpful to distinguish high-grade from low-grade tumors.^29^ Wongi’s study confirmed that solid tumor (CTR = 100%) and tumor size (cut-off value of 10 mm) were the independent predicting factors for high-grade tumors. Similarly to Wongi’s study, Ryo also suggested that solid nodule (CTR = 100%) was an independent predictor of grade 3 tumors. However, in the present study, we found the tumor size was useless to differentiate the IASLC grade 3 tumors from grade 1 or 2. Although solid tumor on CT was found to be significant between IASLC grade 3 tumors and grade 1 or 2 tumors on univariate analysis, it was not found to be the independent influencing factors for predicting IASLC grade 3 tumors on multivariate analysis. In the present study, we identified two independent factors, the SUV_max_ and the air bronchogram sign or vacuole sign of the lesion, for predicting IASLC grade 3 tumor. IASLC grade 3 tumor showed a significantly higher SUV_max_ than that of grade 1 or 2 tumor [8.61 (4.71, 12.90) *vs.* 2.50 (1.30, 5.10), U = 2077.5, *P <* 0.001]. On the contrary, more IASLC grade 1 or 2 tumors presented with air bronchogram sign or vacuole sign on CT (*P* < 0.05). The different biological characteristics of different grade tumors can explain the usefulness of the above two parameters for prediction. A large number of studies have proven that higher grade tumor often have a higher glucose metabolism,^30,31^ which can not only provide more carbon sources to synthesize more organelles to support the proliferation of high grade tumor, but also to meet the energy demand for high activity and invasion of these tumors.^32^ On the contrary, it is well known that lung adenocarcinoma originates from alveoli and bronchioles and they often make regional bronchioles and alveoli to dilate during the growth process. The attenuation of IASLC grade 1 or 2 tumor is usually low and the dilated bronchioles and alveoli can be easily found on CT.^33^ However, IASLC grade 3 tumors often grow rapidly, which compress the dilated bronchus to make it narrow or collapse. Meanwhile, due to its high proliferation and substantial tumor cells, the tumor often has a high attenuation. Therefore, bronchioles and alveoli sign are usually difficult to find on CT for IASLC grade 3 tumors.^34^

Several studies had attempted to establish the models for preoperative predicting IASLC grade 3 tumors. A predicting model built by Min was established mainly on CT signs, while that built by Wongi was based on CT signs and clinical information, such as age and overweight. Although both models showed useful for predicting IASLC grade 3 tumors to some extent, PET metabolic data was not included. As an important parameter that is closely related with tumor aggressiveness, we hypothesis that a model included this parameter may be more helpful to predict the IASLC grade 3 tumors. In the present study, not only more clinical characteristics (sex, age, history of smoking, and tumor markers) and more CT features (tumor diameter, lesion attenuation and some signs) were included to build the model, but also PET metabolic parameters, such as SUV_max_, MTV and TLG, were included, which may make our model can more comprehensively reflect the biological characteristics of tumors and help to predict IASLC grade 3 tumor more accurately. In fact, the present study showed that our model had a high predictive performance with an AUC of 0.825. More importantly, it also had a high negative predictive value of 0.924, which is helpful to accurately rule out IASLC grade 3 tumor 30.

There are some limitations in the present study. Firstly, although a totality of 227 patients were enrolled in present study, a larger sample size would help to get a more reliable results. Secondly, our study was only a single center research; multicenter study would make the model more extensive adaptability. Thirdly, due to shortcoming of the retrospective study, such as case selection bias, a prospective study is warranted. Fourthly, since not all patients with lung cancer undergo PET/CT in the hospital, which may also bring a selection bias.

In summary, the present study reveals that IASLC grade 3 tumors have different clinical, CT and PET characteristics differ from grade 1 or 2 tumor. Compared with grade 1 or 2 tumor, IASLC grade 3 tumors were prone to have mediastinal lymph node metastasis. This tumor often occurs in the patients with male, older age, smoking and increased serum CYFRA21 - 1, CEA, and SCC, and the lesion often presents with solid nodule, burr sign, higher CTR, higher SUV_max_ and TLG. SUV_max_ on PET and air bronchogram sign or vacuole sign on CT are two independent influencing factors for predicting IASLC grade 3 tumor. The prediction model based on these two factors has a high preoperative predictive performance, which may help guide the surgeon to choose a more effective treatment for this highly invasive tumor. A multicenter prospective study is warranted to further confirm our results.

## Conclusion

Our study demonstrates that grade 3 IAC has a unique PET/CT imaging feature. The prediction model established with SUVmax and air bronchogram sign or vacuole sign can effectively predict grade 3 tumors before the operation.

## Data Availability

All data produced in the present study are available upon reasonable request to the authors.

## Abbreviations

IAC: invasive adenocarcinoma
ROC: receiver operating characteristic
SUV_max_: maximum standardized uptake value
IASLC: the International Association for the study of lung cancer
^18^F-FDG: ^18^F-fluorodeoxyglucose
PET/CT: positron emission tomography/computed tomography
ROI: region of interest
MTV: metabolic tumor volume
TLG: and total lesion glycolysis
pGGN: pure ground glass nodule
PACS: picture archiving and communications system
CYFRA21 - 1: cytokeratin fragment 21 - 1
CEA: carcinoembryonic antigen
SCC: squamous cell carcinoma antigen
NSE: neuron-specific enolase
CTR: consolidation-tumor-ratio
RFP: recurrence - free probability
OSEM: ordered subsets expectation maximization
SUV: standardized uptake value
CT: computed tomography
PET: positron emission tomography
[M (Q25, Q75)]: median (25% quantile, 75% quantile)

## Acknowledgments

We thank our colleagues in the Nanfang PET Center who manufactured the radiopharmaceuticals and performed the PET/CT scans.

## References

1. Ettinger DS, Wood DE, Aisner DL, et al. NCCN Guidelines Insights: Non-Small Cell Lung Cancer, Version 2.2021. J Natl Compr Canc Netw. 2021;19:254–266.

2. Siegel RL, Miller KD, Wagle NS, et al. Cancer statistics, 2023. CA Cancer J Clin. 2023;73:17–48.

3. Ettinger DS, Wood DE, Aisner DL, et al. NCCN Guidelines® Insights: Non-Small Cell Lung Cancer, Version 2.2023. J Natl Compr Canc Netw. 2023;21:340–350.

4. Fujikawa R, Muraoka Y, Kashima J, et al. Clinicopathologic and Genotypic Features of Lung Adenocarcinoma Characterized by the International Association for the Study of Lung Cancer Grading System. J Thorac Oncol. 2022;17:700–707.

5. Hill W, Lim EL, Weeden CE, et al. Lung adenocarcinoma promotion by air pollutants. Nature. 2023;616:159–167.

6. Moreira AL, Ocampo PSS, Xia Y, et al. A Grading System for Invasive Pulmonary Adenocarcinoma: A Proposal From the International Association for the Study of Lung Cancer Pathology Committee. J Thorac Oncol. 2020;15:1599–1610.

7. Karasaki T, Moore DA, Veeriah S, et al. Evolutionary characterization of lung adenocarcinoma morphology in TRACERx. Nat Med. 2023;29:833–845.

8. Rokutan-Kurata M, Yoshizawa A, Ueno K, et al. Validation Study of the International Association for the Study of Lung Cancer Histologic Grading System of Invasive Lung Adenocarcinoma. J Thorac Oncol. 2021;16:1753–1758.

9. Weng CF, Huang CJ, Huang SH, et al. New International Association for the Study of Lung Cancer (IASLC) Pathology Committee Grading System for the Prognostic Outcome of Advanced Lung Adenocarcinoma. Cancers (Basel). 2020;12.

10. Yanagawa N, Sugai M, Shikanai S, et al. The new IASLC grading system for invasive non-mucinous lung adenocarcinoma is a more useful indicator of patient survival compared with previous grading systems. J Surg Oncol. 2023;127:174–182.

11. Xu L, Su H, Hou L, et al. The IASLC Proposed Grading System Accurately Predicts Prognosis and Mediastinal Nodal Metastasis in Patients With Clinical Stage I Lung Adenocarcinoma. Am J Surg Pathol. 2022;46:1633–1641.

12. Yoshida C, Yokomise H, Ibuki E, et al. High-grade tumor classified by new system is a prognostic predictor in resected lung adenocarcinoma. Gen Thorac Cardiovasc Surg. 2022;70:455–462.

13. Woo W, Cha YJ, Kim BJ, et al. Validation Study of New IASLC Histology Grading System in Stage I Non-Mucinous Adenocarcinoma Comparing With Minimally Invasive Adenocarcinoma. Clin Lung Cancer. 2022;23:e435–e442.

14. Hou L, Wang T, Chen D, et al. Prognostic and predictive value of the newly proposed grading system of invasive pulmonary adenocarcinoma in Chinese patients: a retrospective multicohort study. Mod Pathol. 2022;35:749–756.

15. Daly ME, Singh N, Ismaila N, et al. Management of Stage III Non-Small-Cell Lung Cancer: ASCO Guideline. J Clin Oncol. 2022;40:1356–1384.

16. Thai AA, Solomon BJ, Sequist LV, et al. Lung cancer. Lancet. 2021;398:535–554.

17. Marulli G, Faccioli E, Mammana M, et al. Predictors of nodal upstaging in patients with cT1-3N0 non-small cell lung cancer (NSCLC): results from the Italian VATS Group Registry. Surg Today. 2020;50:711–718.

18. Zhou Y, Du J, Ma C, et al. Mathematical models for intraoperative prediction of metastasis to regional lymph nodes in patients with clinical stage I non-small cell lung cancer. Medicine (Baltimore). 2022;101:e30362.

19. Kagna O, Solomonov A, Keidar Z, et al. The value of FDG-PET/CT in assessing single pulmonary nodules in patients at high risk of lung cancer. Eur J Nucl Med Mol Imaging. 2009;36:997–1004.

20. Suárez-Piñera M, Belda-Sanchis J, Taus A, et al. FDG PET-CT SUVmax and IASLC/ATS/ERS histologic classification: a new profile of lung adenocarcinoma with prognostic value. Am J Nucl Med Mol Imaging. 2018;8:100–109.

21. Önner H, Coşkun N, Erol M, et al. The Role of Histogram-Based Textural Analysis of (18)F-FDG PET/CT in Evaluating Tumor Heterogeneity and Predicting the Prognosis of Invasive Lung Adenocarcinoma. Mol Imaging Radionucl Ther. 2022;31:33–41.

22. Groheux D, Quere G, Blanc E, et al. FDG PET-CT for solitary pulmonary nodule and lung cancer: Literature review. Diagn Interv Imaging. 2016;97:1003–1017.

23. Sibille L, Seifert R, Avramovic N, et al. (18)F-FDG PET/CT Uptake Classification in Lymphoma and Lung Cancer by Using Deep Convolutional Neural Networks. Radiology. 2020;294:445–452.

24. Miao H, Shaolei L, Nan L, et al. Occult mediastinal lymph node metastasis in FDG-PET/CT node-negative lung adenocarcinoma patients: Risk factors and histopathological study. Thorac Cancer. 2019;10:1453–1460.

25. Huang Z, Wu Y, Fu F, et al. Parametric image generation with the uEXPLORER total-body PET/CT system through deep learning. Eur J Nucl Med Mol Imaging. 2022;49:2482–2492.

26. Zhang X, Qiao W, Kang Z, et al. CT Features of Stage IA Invasive Mucinous Adenocarcinoma of the Lung and Establishment of a Prediction Model. Int J Gen Med. 2022;15:5455–5463.

27. Dong H, Wang X, Qiu Y, et al. Establishment and visualization of a model based on high-resolution CT qualitative and quantitative features for prediction of micropapillary or solid components in invasive lung adenocarcinoma. J Cancer Res Clin Oncol. 2023;149:10519–10530.

28. Woo W, Cha YJ, Park CH, et al. Predictive scoring of high-grade histology among early-stage lung cancer patients: The MOSS score. Thorac Cancer. 2023;14:1865–1873.

29. Liang M, Tang W, Tan F, et al. Preoperative prognostic prediction for stage I lung adenocarcinomas: Impact of the computed tomography features associated with the new histological grading system. Front Oncol. 2023;13:1103269.

30. Sun XY, Chen TX, Chang C, et al. SUVmax of (18)FDG PET/CT Predicts Histological Grade of Lung Adenocarcinoma. Acad Radiol. 2021;28:49–57.

31. Nakamura H, Saji H, Shinmyo T, et al. Close association of IASLC/ATS/ERS lung adenocarcinoma subtypes with glucose-uptake in positron emission tomography. Lung Cancer. 2015;87:28–33.

32. Bu L, Tu N, Wang K, et al. Relationship between (18)F-FDG PET/CT Semi-Quantitative Parameters and International Association for the Study of Lung Cancer, American Thoracic Society/European Respiratory Society Classification in Lung Adenocarcinomas. Korean J Radiol. 2022;23:112–123.

33. Kitazawa S, Saeki Y, Kobayashi N, et al. Three-dimensional mean CT attenuation value of pure and part-solid ground-glass lung nodules may predict invasiveness in early adenocarcinoma. Clin Radiol. 2019;74:944–949.

34. Zheng X, Lin J, Xie J, et al. Evaluation of recurrence risk for patients with stage I invasive lung adenocarcinoma manifesting as solid nodules based on (18)F-FDG PET/CT, imaging signs, and clinicopathological features. EJNMMI Res. 2023;13:52.

