## Supplementary Materials for "^18^F-FDG PET/CT characteristics of IASLC grade 3 invasive adenocarcinoma and the value of ^18^F-FDG PET/CT for preoperative prediction"

**Supplement materials**

1. **Acquisition for ^18^F-FDG PET/CT**

The patients were instructed to be fast for more than 6 hours, and the blood glucose was measured before ^18^F-FDG injection to ensure that the blood glucose was within 11.1 mmol/L. For patients with diabetes, the blood glucose should be controlled below 11.1 mmol/L before ^18^F-FDG injection. ^18^F-FDG was administrated intravenously with a dose of 259~444 Mbq (7.0 - 12 mCi, 150 uCi/kg). PET/CT scan was performed at approximately 60 min after ^18^F-FDG injection. Multiple bed-positions scan with 3D 2 min/bed position was acquired for Biograph mCTx, which covered the scope from the cranial roof to the upper femur and single bed-position with 5 min was acquired for uEXPLORER which covered the total body. CT parameters: voltage 120 kV for both Biograph mCTx and uEXPLORER PET/CT, current automatic milliampere for also both Biograph mCTx and uEXPLORER PET/CT. PET images were reconstructed using the ordered subsets expectation maximization (OSEM) and CT was reconstructed with standard soft and lung method. The PET and CT images were transferred to the workstation for image alignment fusion, and the CT data were used for attenuation calibration for the PET images.

2. **Thin-section acquisition**

A limited thin-section CT was performed for the lung nodules with single-breath-holding, which only covered a limited field of the lung nodule, using the CT scanner of PET/CT. This regional thin-section CT provides a high-quality image to evaluate the nodule but exposes the patients to only limited radioactivity. The acquisition parameters were 140 kVp, 160 mA, a pitch of 0.875, and 0.625 mm collimation. No intravenous contrast was used. After scanning, the thin-section CT images were reconstructed into 1.0-mm-thick sections using high-frequency algorithms.
